# Quantifying Blood Culture Volume Using an Automated System: Insights from Pediatric and Adult Simulated Collections Using BACTEC FXI

**DOI:** 10.64898/2026.08.21.26361057

**Authors:** David Turner, Joshua Herr

## Abstract

**Objectives:** Capturing adequate blood volume for blood cultures is critical for accurate detection of bloodstream infections. Pediatric volume targets vary by age and weight, whereas adult targets are standardized. The BD BACTEC™ FXI Culture System (FXI) contains an integrated calibrated load cell capable of automatically reporting blood volume measurements for each vial loaded onto the system. This study evaluated the accuracy of the FXI”s blood volume measurements in simulated pediatric and adult patients.

**Methods:** Mock pediatric and adult blood draws were performed, using bagged whole blood, to replicate real-world collection protocols. Syringe-collected blood volumes ranged from 2.0 to 15.0 mL for pediatric patients, depending on mock patient weight, and were fixed at 40.0 mL for adults. Samples were inoculated into BD BACTEC™ Peds Plus™/F, Plus Aerobic/F, and Lytic/10 Anaerobic/F Culture Vials, with a target volume of 2.0 to 10.0 mL per bottle. Reference blood volumes were determined gravimetrically using manually obtained pre- and post-inoculation weights with a blood-specific gravity of 1.055 g/mL and were compared to the automatically measured, gravimetric-based blood volumes reported by the BACTEC™ FXI Culture System.

**Results:** Automated volume estimates were accurate to a mean error of –0.03 mL per bottle (SD, 0.40 mL; n=168; 95% CI, -0.09 mL, 0.03 mL) and –0.08 mL (SD, 0.79 mL; n=72; 95% CI, -0.26 mL, 0.10 mL) when assessing total volume collected per patient.

**Conclusions:** Our findings demonstrate that the automated system can quantify blood volumes in BACTEC culture vials and support blood volume monitoring for pediatric and adult collections. The gravimetric approach is also amenable to full automation for efficient and accurate blood volume determination.

## 1. INTRODUCTION

Collecting adequate blood culture volumes is essential to ensure timely detection and identification of blood-borne pathogens and appropriate clinical intervention.(1) The diagnostic yield of blood cultures is influenced by multiple factors, including adherence to clinical collection guidelines, proper handling and transport of blood culture bottles, and ensuring that each bottle contains an adequate volume of blood.(2, 3) Within the scope of the microbiology laboratory workflow, collecting accurate blood volumes in blood culture bottles is essential to ensure reliable blood culture results;(4) accordingly, the Clinical and Laboratory Standards Institute (CLSI) recommends routine monitoring of blood volume as a key part of laboratory workflow process.(5) While visual inspection and manual weighing are conventional methods for assessing bottle fill volume in laboratory workflows, commercial blood culture systems also estimate blood volume using technologies such as optical sensors and blood background fluorescence analysis. However, human factors may impact the accuracy of visual methods and sensitivity of the built-in measurement tools may be affected by factors such as label placement(6) or bottle foaming(7). To address these challenges, the BACTEC FXI Culture System was developed with an integrated calibrated load cell that uses a gravimetric approach to provide automated blood volume measurement for individual bottles upon specimen loading. The system uses the measured weight of inoculated bottles from the integrated load cell and subtracts the established weight of uninoculated bottles to derive the inoculated sample volumes. Our study evaluated the accuracy of this novel system using pediatric and adult blood culture bottles.

## 2. METHODS

This investigation was conducted November 7, 2024 to May 27, 2025 at BD R&D laboratory (Sparks, MD) using bagged whole blood obtained from BioIVT, LLC with donor consent, as approved by Western Copernicus Group Institutional Review Board (WCG IRB #20161665). For the simulated pediatric blood collection, BD BACTEC™ Peds Plus™/F Culture Vials (Becton, Dickinson and Company, Sparks, MD) were first weighed using a calibrated laboratory balance accurate to 0.01 g. Blood culture bottles were then inoculated to simulate pediatric patients using syringe draws of bagged whole blood, following draw protocols for this population(8): ≤3.0 kg (1×2mL), >3.0–5.0 kg (1×3mL), >5.0–7.0 kg (1×5mL), >7.0–12.0 kg (2×5mL), and >12.0–20.0 kg (3×5mL).

For the simulated adult blood collection process, de-capped BD BACTEC™ Plus Aerobic/F and BD BACTEC™ Lytic/10 Anaerobic/F Culture Vials (Becton, Dickinson and Company, Sparks, MD) were weighed using a calibrated analytical laboratory balance accurate to 0.01 g. For this simulated adult population, 10 mL of whole bagged blood was injected by syringe into four bottles—two aerobic and two anaerobic—representing a total draw of 40 mL per mock patient.

Post-inoculation reference measurement was performed by weighing all bottles (pediatric and adult), assessing the difference between bottle weight before vs after inoculation, and dividing the weight difference by the specific gravity of blood (1.055 g/mL). For blood volume measurement using the automated BD BACTEC™ FXI Culture System (Becton, Dickinson and Company, Sparks, MD), the same blood culture bottles were automatically weighed on input using an integrated calibrated load cell. Media product specific pre-inoculation weights, obtained from manufacturing data, were subtracted from filled bottle weights to calculate blood volume.

A custom software application corrected for factors such as load cell zero value and automatically converted results to a volumetric output. Reference blood volumes were subsequently compared to the automatically measured blood volumes reported by the BACTEC FXI Culture System.

## 3. RESULTS

A total of 72 blood collections were performed using bagged blood to represent 50 simulated pediatric patients across five weight categories, and 22 simulated adult patients (Table 1). These 72 blood collections yielded 168 inoculated blood culture bottles. For simulated pediatric patients, the blood volume was distributed among one to three culture bottles, resulting in a total of 80 BACTEC Peds Plus Culture Vials. For simulated adult patients, 40 mL of blood was collected per patient and equally distributed among four culture bottles (two BACTEC Plus Aerobic/F Vials and two BACTEC Lytic/10 Anaerobic/F Vials) resulting in 44 bottles of each type for a total of 88 bottles.

**Table 1.**
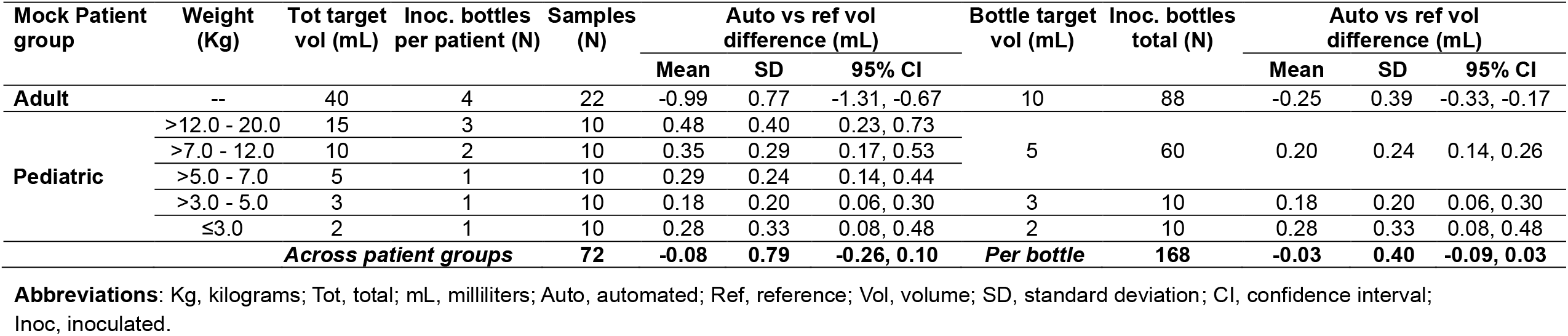
Automated vs. Reference Volume Error Across Patient Weight Groups and Bottle Target Volumes.

Compared to the reference method, the automated calculation of blood volume per bottle demonstrated high accuracy, with a mean error of -0.03 mL (SD, 0.40 mL; n = 168; 95% CI, -0.09 mL, 0.03 mL). Similarly, for total blood volume collected per simulated patient, the mean error was -0.08 mL (SD, 0.79 mL; n = 72; 95% CI, -0.26 mL, 0.10 mL) (Table 1). Automated blood volume calculations appeared closely aligned with reference volumes across all pediatric weight categories and adult collections (Figure 1). No statistically significant difference was found between the automated blood volume measurements and the reference volumes using a paired t-test (*p* = 0.287).

**Figure 1.**
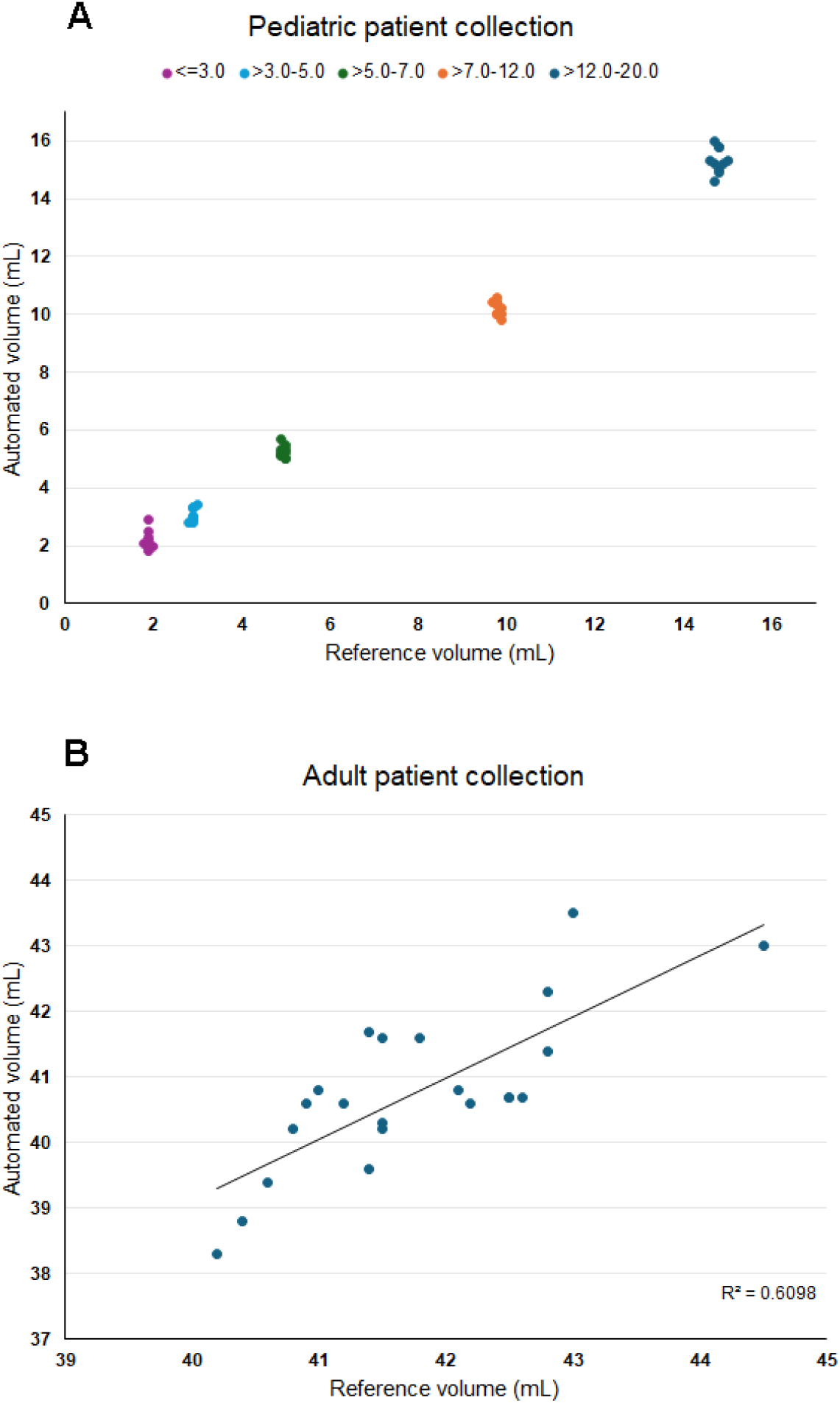
Agreement between automated and reference volume measurements in pediatric and adult simulated blood collections. **(A)** represents automated versus reference volumes for simulated pediatric patient collections across five reference-volume ranges. Each data point represents an individual collection color-coded by reference-volume category. **(B)** shows a scatterplot of automated versus reference volumes for simulated adult patient collections (40 mL total target volume).

## 4. DISCUSSION

Our study demonstrates the level of accuracy that can be achieved using an automated, gravimetric blood volume measuring system in both adult and pediatric populations. When reference blood volumes were compared to those calculated by the automated system, a mean error of -0.03 mL per bottle and -0.08 mL per simulated patient were observed, representing minimal deviations from expected values, and suggesting strong reliability for automated volume monitoring in blood culture. The mean error ranged from -0.25 mL for bottles containing 10 mL of blood to 0.28 mL for bottles containing 2 mL, indicating that gravimetric blood volume measurement is accurate across the range of blood volumes typically encountered in clinical laboratories, including suboptimal adult blood culture volumes (<8 mL). These results suggest a more favorable performance of our automated blood volume measurement system compared to that of BacT/ALERT® VIRTUO (VIRTUO; bioMérieux, Marcy l”Étoile, France). Using VIRTUO, a study by Lee and Kim reported height-based optical measurements with median differences of 0.21 and 1.40 mL, for anaerobic and aerobic bottles, respectively, compared to weight-based measurement.(9) More recently, Kim and Park added to these findings using an updated VIRTUO software, reporting that height-based measurements tended to overestimate blood volume by a mean difference of 1.29 and 0.05 mL for anaerobic and aerobic bottles, respectively, relative to gravimetric reference measurements.(7) Together, these data suggest that gravimetric measurement strategies may provide improved accuracy and consistency across bottle types and clinical use cases when compared with height- or blood background-based methods.

While the findings of this study demonstrate the potential benefits of automated blood volume measurement, some limitations should be acknowledged. First, the sample size was limited, which may affect the generalizability of the results. Additionally, the population from which blood was initially collected may not fully represent the diversity of patients encountered in clinical practice.

Beyond analytical performance, adoption of fully automated gravimetric blood volume measurement systems may offer meaningful clinical and operational benefits. Most critically, having an immediate record of blood volume collected in each culture bottle helps laboratories determine the actual volume delivered to each bottle, which is an important factor to confirm the presence of a bloodborne infection. Further, accurate, automated volume verification supports compliance with CLSI recommendations for routine blood volume monitoring while reducing reliance on manual weighing or subjective visual inspection. From a laboratory workflow perspective, integration of automated blood volume assessment may improve standardization, reduce hands-on time, and enable real-time feedback to clinical teams regarding suboptimal collections, thereby supporting improved diagnostic yield and more reliable interpretation of negative blood culture results.

Our findings indicate that the automated system can accurately quantify blood volumes in BACTEC Peds Plus/F, Plus Aerobic/F, and Lytic/10 Anaerobic/F Culture Vials, enabling reliable monitoring for pediatric and adult samples. The gravimetric method has been incorporated into the BACTEC FXI Culture System as a fully automated solution to support accurate and efficient volume determination.

## Data Availability

Data will be made available upon reasonable request to the corresponding author.

## DECLARATION OF GENERATIVE AI AND AI-ASSISTED TECHNOLOGIES IN THE MANUSCRIPT PREPARATION PROCESS

During the preparation of this manuscript, Microsoft 365 Copilot Business was used to improve the clarity of selected sentences that were initially human generated. Artificial intelligence was not used to generate, develop, or modify tables or figures, nor was it used for data analysis. Following the use of this tool, the authors reviewed, edited, and approved all content as necessary and take full responsibility for the accuracy and integrity of the published article.

## ACKNOWLEDGEMENTS

Thank you to Christopher Massey, Becky Newcomb, and Kristina Robinson from Waters Advanced Diagnostics (formerly BD Diagnostic Solutions) for their contribution to the study. Thank you also to Hélène Tanguay, PhD, and Brian G. Vassallo, PhD, for medical writing assistance, and Dorsey Mills, BA, for editorial assistance. All are employees of Waters Advanced Diagnostics (formerly BD Diagnostic Solutions). Portions of this work were previously presented in abstract and poster form at scientific conferences, including the American Society for Microbiology (ASM 2023-2025) and the European Society of Clinical Microbiology and Infectious Diseases (ESCMID 2023-2025).

## AUTHORS CONTRIBUTIONS

DT: Conceptualization, Methodology, Formal Analysis, Data Curation, Writing – Original Draft, Manuscript Review & Editing, Visualization.

JH: Conceptualization, Methodology, Manuscript Review & Editing, Supervision, Visualization, Project administration, Funding acquisition.

## FUNDING

This study was funded by Becton, Dickinson and Company (now Waters Advanced Diagnostics).

## POTENTIAL CONFLICTS OF INTEREST

DT and JH are employees of Waters Advanced Diagnostics (formerly BD Diagnostic Solutions) and own BD/Waters shares.

## Notes

### Author Declarations

Western Copernicus Group Institutional Review Board (WCG IRB #20161665) gave ethical approval for this work.

## REFERENCES

1. Lamy B, Dargère S, Arendrup MC, Parienti JJ, Tattevin P. How to Optimize the Use of Blood Cultures for the Diagnosis of Bloodstream Infections? A State-of-the Art. Front Microbiol. 2016;7:697. doi:10.3389/fmicb.2016.00697.

2. Mermel LA, Maki DG. Detection of bacteremia in adults: consequences of culturing an inadequate volume of blood. Ann Intern Med. 1993;119(4):270–2. doi:10.7326/0003-4819-119-4-199308150-00003.

3. Deas G, Hamilton F, Williams P. Less haste, more speed: Does delayed blood culture transport time lead to adverse incubation times or yield? Journal of Infection. 2025;91(1). doi:10.1016/j.jinf.2025.106520.

4. Cattoir L, Claessens J, Cartuyvels R, Van den Abeele AM. How to achieve accurate blood culture volumes: the BD BACTEC FX blood volume monitoring system as a measuring instrument and educational tool. Eur J Clin Microbiol Infect Dis. 2018;37(9):1621–6. doi:10.1007/s10096-018-3291-x.

5. CLSI. Principles and Procedures for Blood Cultures. 2nd ed. CLSI guideline M47. Clinical and Laboratory Standards Institute; 2022. Retrieved Oct. 22, 2025: https://www.standards-global.com/wp-content/uploads/pdfs/preview/2253411.

6. Emeraud C, Yilmaz S, Fortineau N, Cuzon G, Dortet L. Quality indicators for blood culture: 1 year of monitoring with BacT/Alert Virtuo at a French hospital. J Med Microbiol. 2021;70(3). doi:10.1099/jmm.0.001300.

7. Kim K, Park S. Validation of the Accuracy of Automatic Measurement of Blood Volume in Culture Bottles for Blood Culture. Diagnostics (Basel). 2023;13(16). doi:10.3390/diagnostics13162685.

8. Huber S, Hetzer B, Crazzolara R, Orth-Holler D. The correct blood volume for paediatric blood cultures: a conundrum? Clin Microbiol Infect. 2020;26(2):168–73. doi:10.1016/j.cmi.2019.10.006.

9. Lee S, Kim S. Accuracy of BacT/Alert Virtuo for Measuring Blood Volume for Blood Culture. Ann Lab Med. 2019;39(6):590–2. doi:10.3343/alm.2019.39.6.590.

